# The Rapid Assessment of Aggregated Wastewater Samples for Genomic Surveillance of SARS-CoV-2 on a City-Wide Scale

**DOI:** 10.1101/2021.08.17.21262170

**Authors:** Eric C. Rouchka, Julia L. Chariker, Kumar Saurabh, Sabine Waigel, Wolfgang Zacharias, Mei Zhang, Daymond Talley, Ian Santisteban, Madeline Puccio, Sarah Moyer, Rochelle H. Holm, Ray A. Yeager, Kevin Sokoloski, Joshua Fuqua, Aruni Bhatnagar, Ted Smith

## Abstract

Throughout the course of the ongoing SARS-CoV-2 pandemic there has been a need for approaches that enable rapid monitoring of public health using an unbiased and minimally invasive means. A major way this has been accomplished is through the regular assessment of wastewater samples by qRT-PCR to detect the prevalence of viral nucleic acid with respect to time and location. Further expansion of SARS-CoV-2 wastewater monitoring efforts to include the detection of variants of interest / concern through next-generation sequencing have enhanced the understanding of the SARS-CoV-2 outbreak. In this report we detail the results of a collaborative effort between public health and metropolitan wastewater management authorities and the University of Louisville to monitor the SARS-CoV-2 pandemic through the monitoring of aggregate wastewater samples over a period of 28 weeks. Our data indicates that wastewater monitoring of water quality treatment centers and smaller neighborhood-scale catchment areas is a viable means by which the prevalence and genetic variation of SARS-CoV-2 within a metropolitan community of approximately one million individuals may be monitored. Importantly, these efforts confirm that regional emergence and spread of variants of interest / concern may be detected as readily in aggregate wastewater samples as compared to the individual wastewater sheds.

## 1. Introduction

The clinical prevalence of Severe Acute Respiratory Syndrome Coronavirus 2 (SARS-CoV-2) in the early stages of the pandemic led to the application of wastewater surveillance over a range of scales-from pooled sources covering large swaths of a community encompassing hundreds of thousands of individuals, to relatively isolated samples consisting of dozens to hundreds of individuals living in congregate settings [1-6]. These efforts were initially designed to monitor the trends of “true” incidence of SARS-CoV-2 infection within the targeted communities, as wastewater sampling represents a non-invasive means of quantitatively assessing the presence of SARS-CoV-2 nucleic acid derived from individuals participating and non-participating in clinical testing efforts [1,7-12]. Thus, the sampling of wastewater represents a means by which SARS-CoV-2 public health may be assessed on a large scale to determine the disease / infection burden [2,8,9,13-17].

The genetic diversity of SARS-CoV-2 has largely been determined via the sequencing of clinical samples obtained from infected individuals; however, changes in the viral genome have been shown to be detectable in wastewater samples [18-23]. Indeed, the presence of specific SARS-CoV-2 variants is often detectable in the wastewater of a community prior to detection in clinical samples [2,5,9,13,21,23,24]. As such, the genomic surveillance of SARS-CoV-2 through wastewater sampling represents a cost-effective and unbiased means by which the community prevalence and genetic diversity of SARS-CoV-2 infections may be assessed.

In this report we describe efforts undertaken by a partnership between the Center for Predictive Medicine and Emerging Infectious Diseases at the University of Louisville, the Kentucky IDeA Networks of Biomedical Research Excellence (KY INBRE), Christina Lee Brown Envirome Institute, the Louisville Metropolitan Sewer District, and the public health department of the city of Louisville, Kentucky to monitor SARS-CoV-2 via the assessment of aggregated wastewater samples. Specifically, our efforts have confirmed the utility and importance of aggregated wastewater surveillance as a means by which to monitor infection rates in a city-wide setting, and we further demonstrate that the emergence of SARS-CoV-2 variants can be readily detected with respect to time and location. In addition to validating the power of wastewater analyses, we also report the detection of several polymorphisms with the likelihood to impact SARS-CoV-2 transmission.

## 2. Results

### 2.1. Detection of SARS-CoV-2 RNA in Aggregate Wastewater Samples Correlates with Community Prevalence

As shown in Figure 1, the weekly incidence of SARS-CoV-2 infection in Louisville KY steadily increased over an approximately 3-month period, corresponding to the date range of October 2020 to January 2021, with peak incidence occurring during the first week of January 2021. During this time the analysis of aggregated wastewater samples obtained from the five Water Quality Treatment Centers (WQTCs) that service the Louisville metropolitan area demonstrated the clear presence of SARS-CoV-2 nucleic acid as detected by qRT-PCR. As expected, increased detection of SARS-CoV-2 RNAs correlated with the observed increases in community prevalence as reflected by the available public health data. Nonetheless, the increases among the two measurements were temporally asynchronous, in that levels of viral RNA in the aggregated wastewater increased approximately one week earlier than the reported increases in incidence as determined via clinical testing. After reaching peak incidence in early January 2021, the weekly incidence of confirmed SARS-CoV-2 steadily decreased to levels below those observed in October 2020. Notably, despite the significant decrease in clinically confirmed SARS-CoV-2 cases over the next 3 months in the Louisville metropolitan area, the detection of SARS-CoV-2 RNA in aggregate wastewater samples remained at levels similar to those observed during the peak incidence period. This discrepancy remained until Week 24, whereafter the levels of SARS-CoV-2 nucleic acid detected via qRT-PCR began to steadily decrease back to levels consistent with those observed before the January 2021 peak.

**Figure 1.**
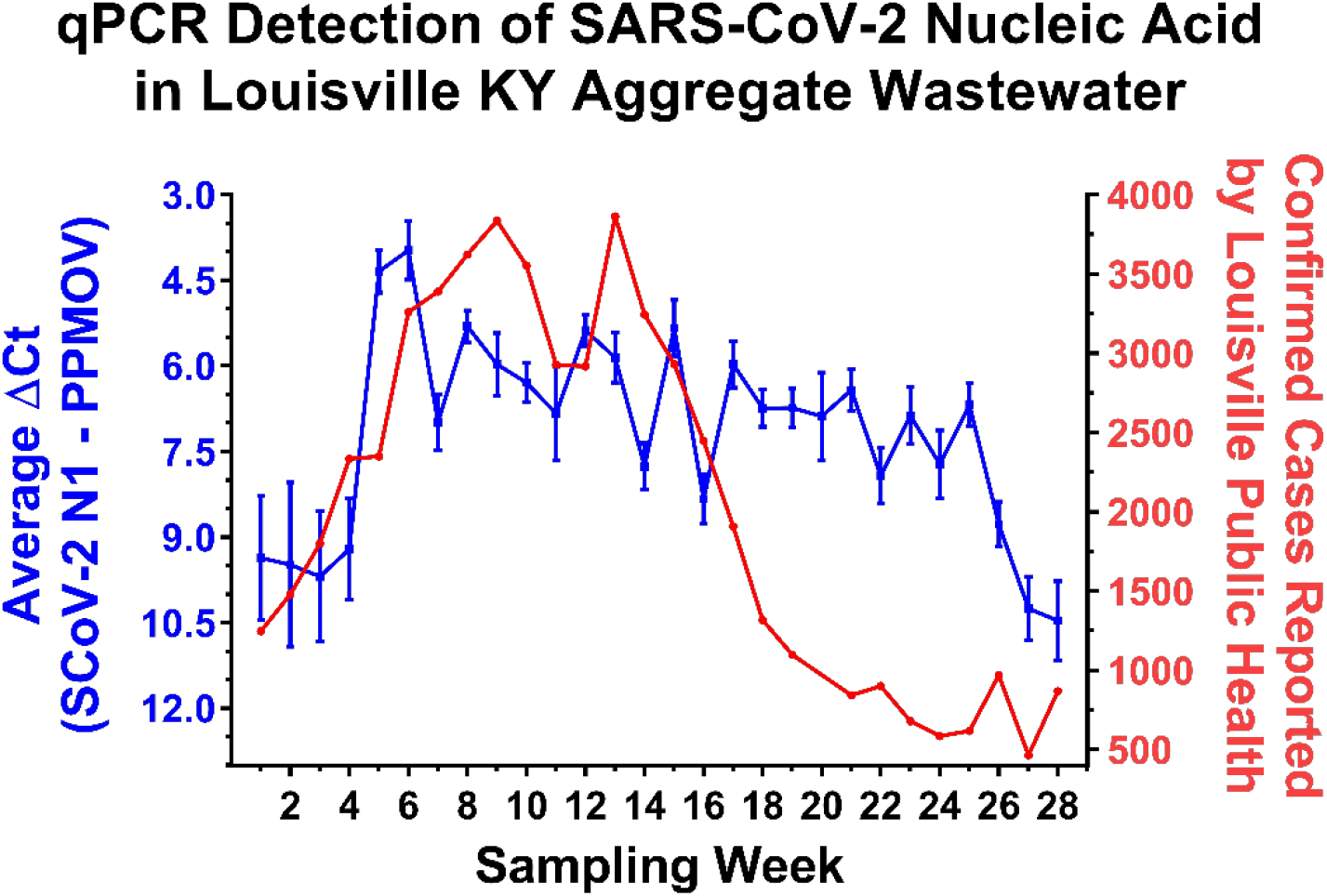
The Quantitative Detection of SARS-CoV-2 in the Aggregated Wastewater of Water Quality Treatement Plants, Louisville, Kentucky. On the left Y-axis (Blue) are the averaged ΔCt values obtained from the 5 water quality treatment centers (WQTCs) that service the Louisville metropolitan area. The ΔCt values represent the levels of SARS-CoV-2 N1 amplicon normalized to the PPMOV control amplicon. Note that the left Y-axis is plotted in an inverse manner. Individual data points represent the means of three technical replicates per WQTC per timepoint, with the error bars representing the standard deviation of the means. Plotted on the right Y-axis (Red) are the numbers of confirmed SARS-CoV-2 cases reported to the city of Louisville public health authorities (as per publically available health data) on a per week basis. The X-axis represents the particular week to which the data belongs, with week 1 pertaining to the first week of October 2020, and week 28 being the last week of April 2021. The week pertaining to that of the 3^rd^ and 4^th^ weeks of December are omitted from the above data set, as the underlying public health and wasewater data sets were incomplete for that period of time. The relevant public health data may be found in the supplemental data files accompanying this manuscript.

The discrepancy between the SARS-CoV-2 nucleic acid detection and clinical incidence data sets could be due to a multitude of factors. Perhaps the most obvious potential reason for the loss of correlation between the public health data and the aggregate wastewater sample data during this period is the undercounting of infected individuals or the possibility of viral shedding that contunues for weeks after symptomatic infection [25]. However, it also remained possible that aggregate wastewater sources exhibit reduced exchange rates relative to smaller wastewater tributaries leading to the “stagnation” of viral RNA levels. If the observations reported above for the aggregate wastewater sources were due to slower rates of exchange, then smaller wastewater sources with higher relative rates of exchange would be expected to exhibit contrary behaviors to the larger aggregate sites. Thus, to determine whether flow rate was impacting the aggregate wastewater observations we utilized qRT-PCR to assay smaller wastewater tributaries that fed into the two major WQTCs of Louisville, Morris Forman WQTC and Derek R. Guthrie WQTC. As exhibited by the data presented in Fig. 2, the trends exhibited by these smaller wastewater tributaries closely match the observations obtained for the larger aggregate WQTCs into which they feed. Similar observations were made across other wastewater tributaries in the Louisville metropolitan area. Additionally, the observations for the smaller wastewater sheds are more or less consistent with the averaged measurements for the entire city of Louisville, as reported in Fig. 1.

**Figure 2.**
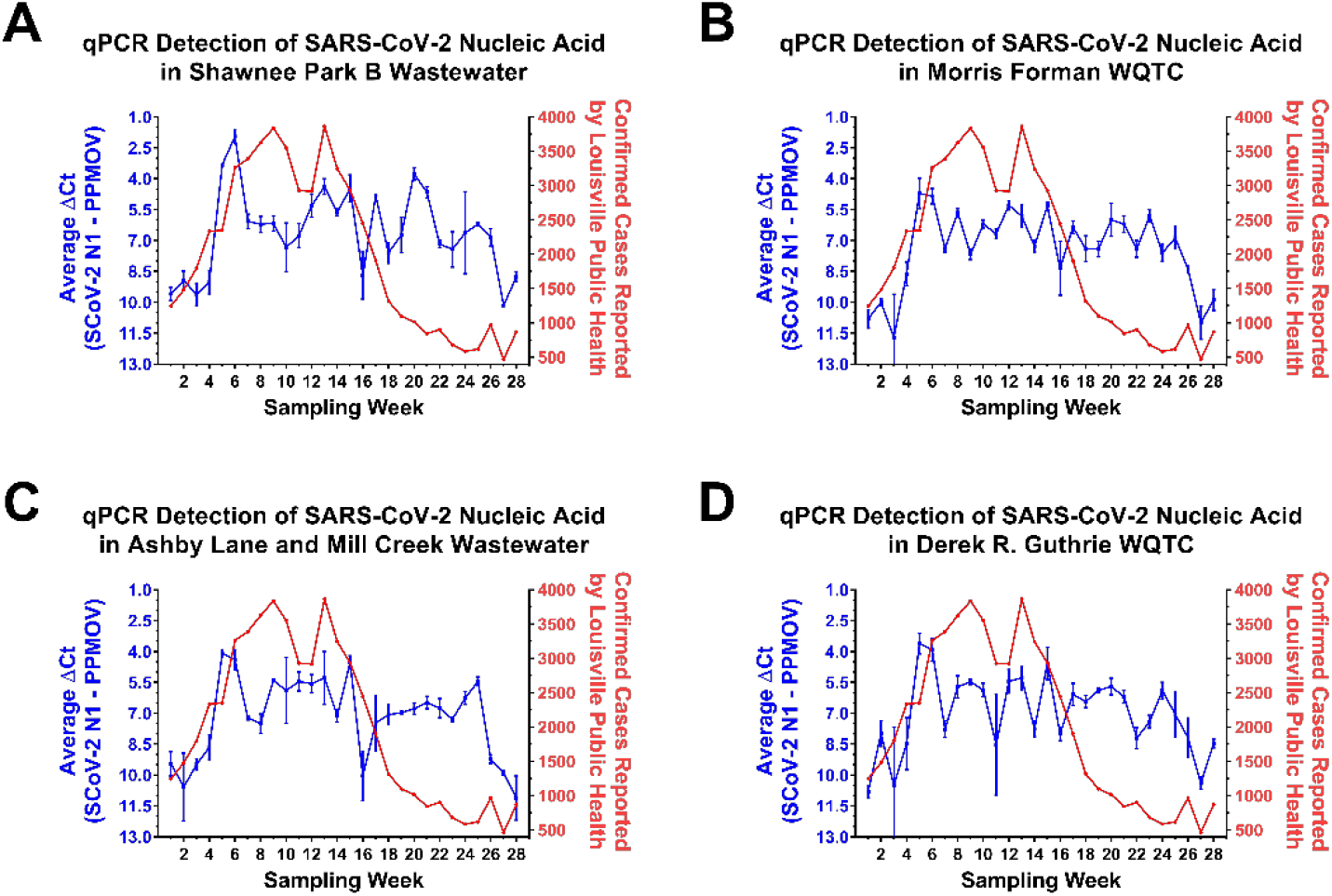
The Quantitative Detection of SARS-CoV-2 in Wastewater Tributaries of the Morris Forman and Derek R. Guthrie WQTCs. On the left Y-axes (Blue) are the average ΔCt values obtained from wastewater tributaries associated with the Shawnee Park B wastewatershed (A), and the aggregate wastewater treatment center to which it feeds-Morris Forman WQTC (B). Panels (C) and (D) are similar, but the areas represented are the Ashby Lane and Mill Creek tributary and the Derek R. Guthrie WQTC. The ΔCt values represent the levels of SARS-CoV-2 N1 amplicon normalized to the PPMOV control amplicon. Note that the left Y-axis is plotted in an inverse manner. Individual data points represent the means of three technical replicates per WQTC per timepoint, with the error bars representing the standard deviation of the means. Plotted on the right Y-axis (Red) are the numbers of confirmed SARS-CoV-2 cases reported to the city of Louisville public health authorities on a per week basis. The X-axis represents the particular week to which the data belongs, with week 1 pertaining to the first week of October 2020, and week 28 being the last week of April 2021. The week pertaining to that of the 3^rd^ and 4^th^ weeks of December are omitted from the above data set, as the underlying data sets were incomplete for that period of time.

Together these observations confirm that the analysis of aggregated wastewater samples obtained from treatment plants are capable of indicating the presence of SARS-CoV-2 nucleic acid. In addition, our analyses of wastewater tributaries and their corresponding individual treatment centers indicate that the detection of SARS-CoV-2 RNA is more or less equitable across local, regional, and metropolitan-wide wastewater sources. While SARS-CoV-2 RNA levels correlated with increasing community prevalence they did not immediately correlate with a reported decrease community incidence, and the likely underlying causes of which are discussed in greater depth later.

### 2.2. Aggregate Wastewater Sources Can Provide a Rapid Means to Detect the Prevalence of Variants of Interest / Concern

While the detection of SARS-CoV-2 RNA levels in aggregate wastewater is useful in determining the presence and prevalence of SARS-CoV-2 within a community, it is also potentially a valuable resource by which the genetic diversity of SARS-CoV-2 in a community may be monitored [18,19,21-24]. To this end, the samples obtained from aggregated wastewater sources during an 8-week period corresponding to the 1st week of March to the final week of April 2021 were subjected to next-generation sequencing to determine the sequence diversity of the circulating SARS-CoV-2 populations. As exhibited in Fig. 3, the depth of coverage across the SARS-CoV-2 genome obtained from the aggregate wastewater samples was more than sufficient to enable the detection of sequence polymorphisms. Moreover, the genome coverage and depth of sequencing correlated with the prevalence of SARS-CoV-2 within the sample (as per comparisons of Figs. 1-3). Unsurpisingly, the depth of coverage for obtained from the sequecing efforts was dependent on the concentration of the sample, as detected by qRT-PCR (Fig. 3A).

**Figure 3.**
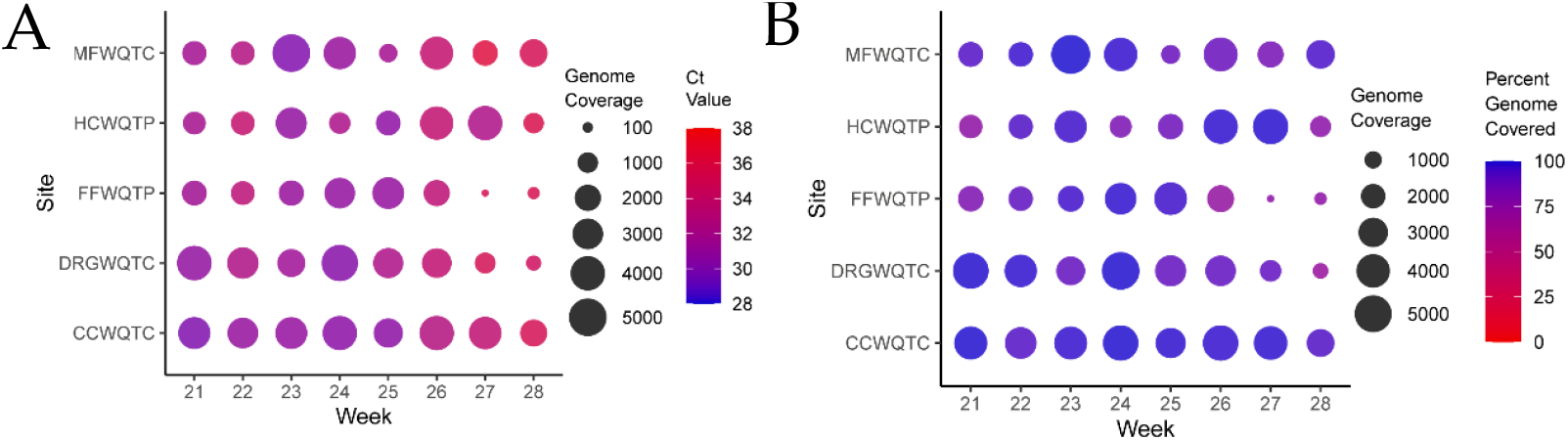
Quantitative Analysis of Per-Base Genome Coverage and Percent Genome Covered for the Aggregate Wastewater Sites. **A)** Shown above is a circle plot depicting the depth of coverage per nucleotide (circle size) and the color of the datapoint corresponds to the Ct value of the associated sample, as detected by qRT-PCR. **B**) Similar to that presented in A, but the circle size represents the depth of coverage, and the color of the circle represents the percentage of the genome covered by at least ten reads (circle color). The individual aggregate wastewater sites are listed on the left vertical axis, and the sampling week listed on the bottom horizontal axis.

The detection of SARS-CoV-2 variants has been a subject of extensive research and public health interest due to their potential impact to global public health. Collectively these efforts have led to the identification of numerous strains with potential consequences to transmission, virulence, and vaccine escapism. At the time of the preparation of this manuscript (May 25th 2021) 4 specific variants have been identified by the CDC as variants of interest (VOIs)- the B.1.526 and B.1.526.1 variants first detected in New York USA, the B.1.525 variant first detected in the United Kingdom and Nigeria, and the P.2 variant first detected in Brazil; and 5 strains have been identified as variants of concern (VOCs)- the B.1.1.7 variant first reported in the United Kingdom, the P.1 variant first detected in Japan but of likely Brazilian origin, the B.1.351 variant first reported in South Africa, the B.1.427 and B.1.429 variants first reported in California USA.

Due to the complex nature of wastewater samples, the isolation and detection of long contiguous sequence reads has been prohibitive. Thus, efforts to identify the genetic composition of the SARS-CoV-2 circulating within populations have utilized the detection of polymorphism markers associated with the VOIs and VOCs. While this approach does not allow for the specific and certain identification of a particular VOI or VOC it allows for the determination of likelihood on a community scale that compliments traditonal clinical survellance strategies, especially when clinical surveillance has a limited scope.

To identify and establish an unbiased threshold of significance by which to gauge the presence or absence of a given genetic marker, the level of variation of laboratory cultured WA1/2020 was assessed in parallel to wastewater samples using next-generation sequencing. From these studies an average SNP noise rate of ∼1% was detected, resulting in the setting of a conservative threshold for the detection of polymorphisms of >5%.

Hence, any sequence polymorphism that meets or exceeds the threshold of 5% is considered to be a genuine detection event. As shown in Figs. 4A and 4C, there are 89 nonsynonymous amino acid polymorphisms associated with one or more of the previously identified VOI/Cs. The detection and correlation of these polymorphism markers enables a broad determination of the circulating SARS-CoV-2 strains; however, it should be noted that this approach loses resolution between strains with close relatedness, such as the B.1429 and B.1.427 VOCs.

**Figure 4.**
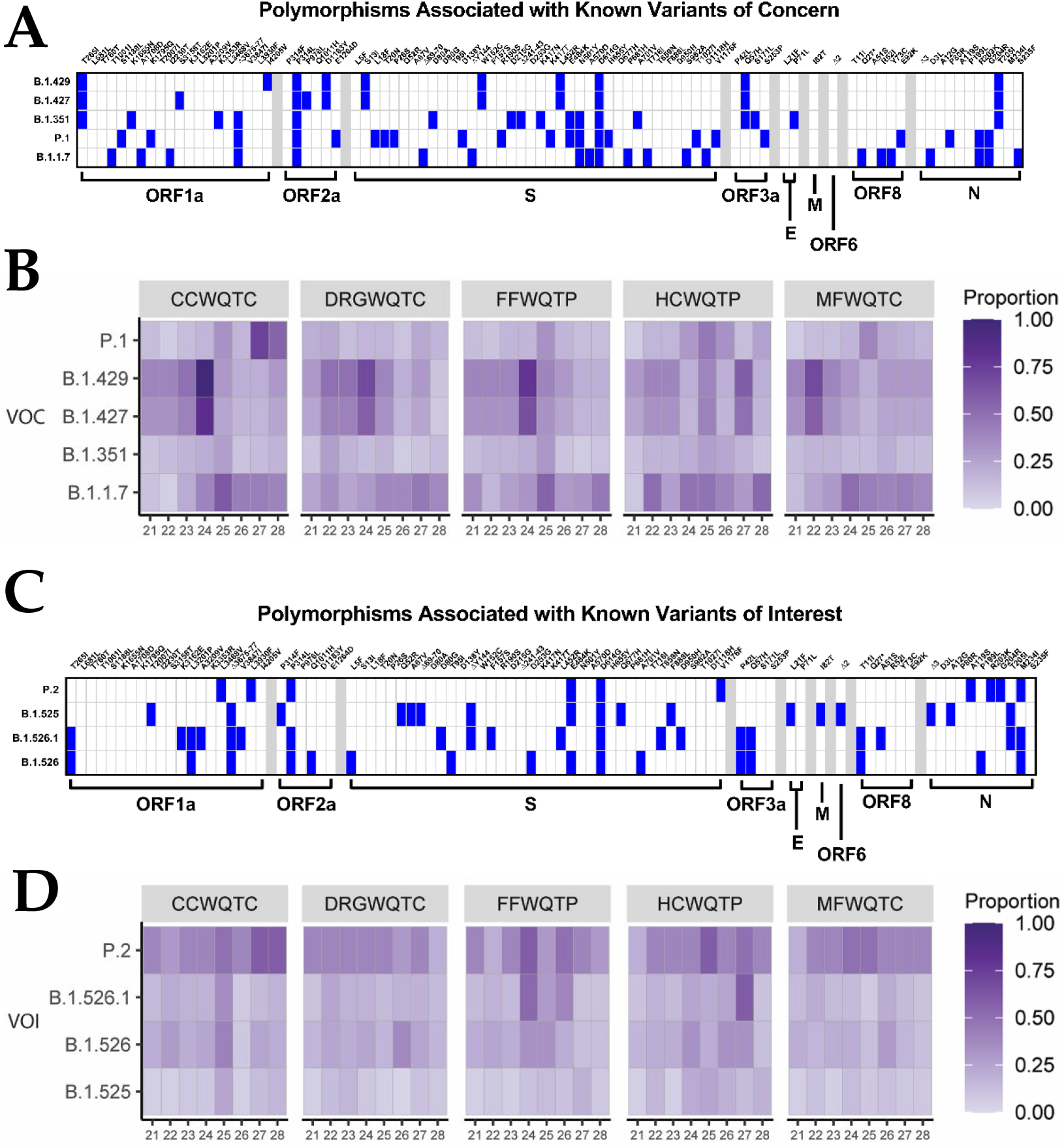
The Identification of Polymorphism Markers Informs About the Presence of Variants of Concern / Interest in the Community. Panels (**A**) and (**C**) detail the specific polymoprhisms associated with each of the currently identified VOCs and VOIs. Specific polymorphisms are found on the upper horizontal axis, and the SARS-CoV-2 gene to which they belong may be found on the lower horizonatal axis. Specific VOC/Is are listed on the left axis. Sequence analyses with respect to time for the aggregated wastewater samples obtained from each of the 5 water quality treatment centers (CC, Cedar Creek; DRG, Derek R. Guthrie; FF, Floyd’s Fork; HC, Hite Creek; and MF, Morris Forman) for each of the identified VOCs (panel B) and VOIs (panel D). Data shown represents the proportion of polymorphism markers which meet or exceed the 5% prevalence threshold that were identified for each listed variant.

Interestingly, the assessment of the aggregated wastewater collection sites via next-generation sequencing revealed that the genetic composition of the community associated SARS-CoV-2 varied significantly during the period of time investigated. Specifically, as depicted in Fig. 4B and 4D, the proportion of polymorphisms associated with a particular VOI or VOC that have met or exceeded the 5% threshold allowed for the determination of which strains were circulating with high likelihood in the community.

A primary observation that may be made from the aggregated wastewater analyses is that the genetic composition of the community associated SARS-CoV-2 of the Louisville metropolitan area varies significantly over a period of several weeks, but is, for the most part, highly consistent across the major wastewater watershed regions within any given week period. Nonetheless, the emergence and expansion of SARS-CoV-2 strains can be detected in the aggregated wastewater sequencing data. This is evidenced by the high prevalence detection of B.1.429 / B.1.427 in the wastewater collected from the MFWQTC at week 4, and the emergence of the B.1.429 / B.1.427 VOCs in the wastewater collected from CCWQTC, DRGWQTC, and FFWQTC. It should be noted that the time period of movement in this instance strongly correlates with the incubation period of SARS-CoV-2.

In addition to detecting the geographic / spatial emergence of the B.1.429 / B.1.427 VOCs, the introduction of new variants to the Louisville metropolitan area was also detected during this time by clinical sampling and aggregate wastewater analysis. In early March 2021 the P.1 variant was identified in a clinical sample taken from an individual in the Louisville metropolitan area on March 5th, 2021. As supported via all available public data, and to the best of our knowledge, this represents the first instance of the P.1 VOC in the state of Kentucky. Contact tracing indicated the likely site of exposure / infection was in Southern Indiana, where the presence of P.1 had recently been detected. During this period of time the markers associated with the P.1 VOC were not detected in the aggregate wastewater of the Louisville metropolitan area indicating likely limited dissemination at that time; however, several weeks later markers associated with the P.1 VOC were detected in the treatment center which services the infected individual’s domicile, MFWQTC. This observation was further confirmed through the analysis of the wastewater tributary associated with the individual’s domicile, namely Shawnee Park B. The local sewershed demonstrated a signal associated with the presence of P1 three weeks before detection in the agreggate WQTC and as with B.1.1.7 predicted the emergence of the variant within the community before it became a stable part of the viral strains present in the population. With the P1 variant, public health efforts including contact tracing were undertaken, but as evidenced by the increased detection of P.1 markers in the Shawnee Park B wastewater samples several weeks after the initial clinical detection of the P.1 VOC, the P.1 variant was not sufficiently contained (Fig. 5). Furthermore, as with the spatial emergence of the B.1.429 / B.1.427 strain, the P.1 strain was later found to have spread to other sites within the city as evidenced by the detection of the P.1 associated VOC markers in the CCWQTC. At this point in time continued spread of the P.1 variant beyond the regions currently identified is expected.

**Figure 5.**
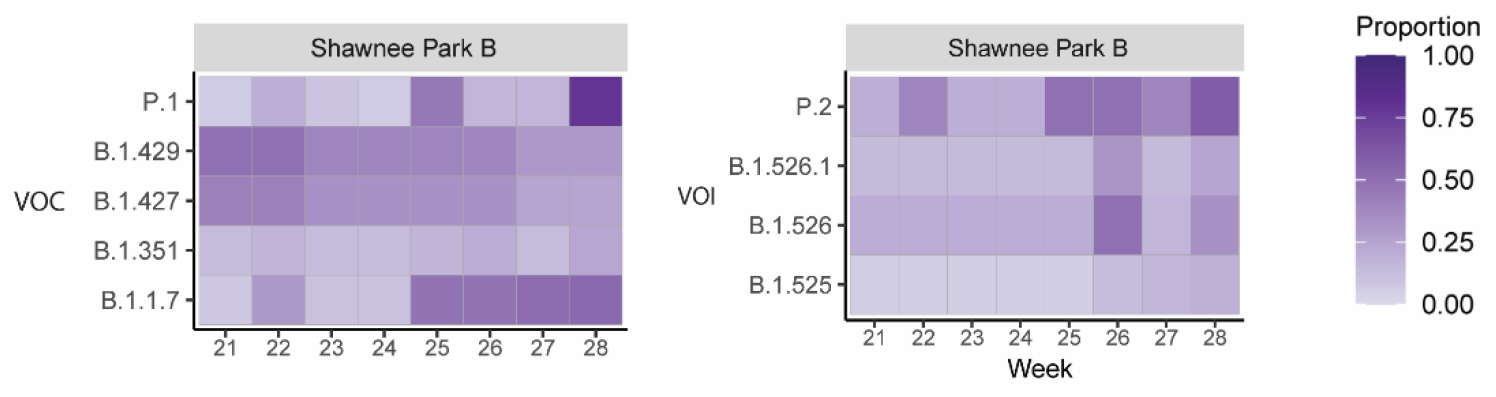
Realtime Identification of the Emergence of the P.1 Variant in a Northwestern Louisville Community. Sequence analyses with respect to time for the wastewater tributary from the Shawnee Park neighborhood in Louisville, KY. Data shown represents the proportion of polymorphism markers which meet or exceed the 5% prevalence threshold that were identified for each listed variant. Week 21 corresponds to the week period / date of the initial detection of the P.1 variant in a clinical sample.

The appearance and emergence of B.1.1.7 is also detected during this period, with the markers associated with this variant becoming prominent concurrent with the rise of B.1.429 / B.1.427, and prior to the emergence of markers associated with the P.1 variant. Whether this variant becomes the dominant strain in the Louisville metropolitan area remains to be seen, but the markers associated with the B.1.1.7 VOC are more prevalent and persist to a higher degree than those for the P.1 VOC over the reported time period.

Altogether these observations confirm that wastewater sampling, even on an aggregated scale covering several hundred thousand persons, is capable of detecting the emergence of variants within a community in real time.

### 2.3. The Analysis of Aggregate Wastewater Sources Can Provide Broad Genetic Information on a Community-Level

As demonstrated above, the analysis of aggregated wastewater allows for rapid broad detection of genetic markers associated with SARS-CoV-2 variants within a community. Further analysis of the sequencing data outside of the lens of known VOI/Cs revealed the presence of other polymorphisms with potential public health ramifications within the Louisville community. As a major focus of SARS-CoV-2 surveillance is potential vaccine escapism we elected to focus our efforts on identifying highly prevalent polymorphisms of the SARS-CoV-2 S protein [26]. As with our previous efforts to detect polymorphisms associated with VOI/Cs, in order to remain conservative in our analyses a prevalence threshold of 5% was established. As shown in Fig. 6, 96 polymorphisms exceeded the threshold of 5% prevalence, however relatively few specific polymorphisms reached saturation or persisted for several weeks.

**Figure 6.**
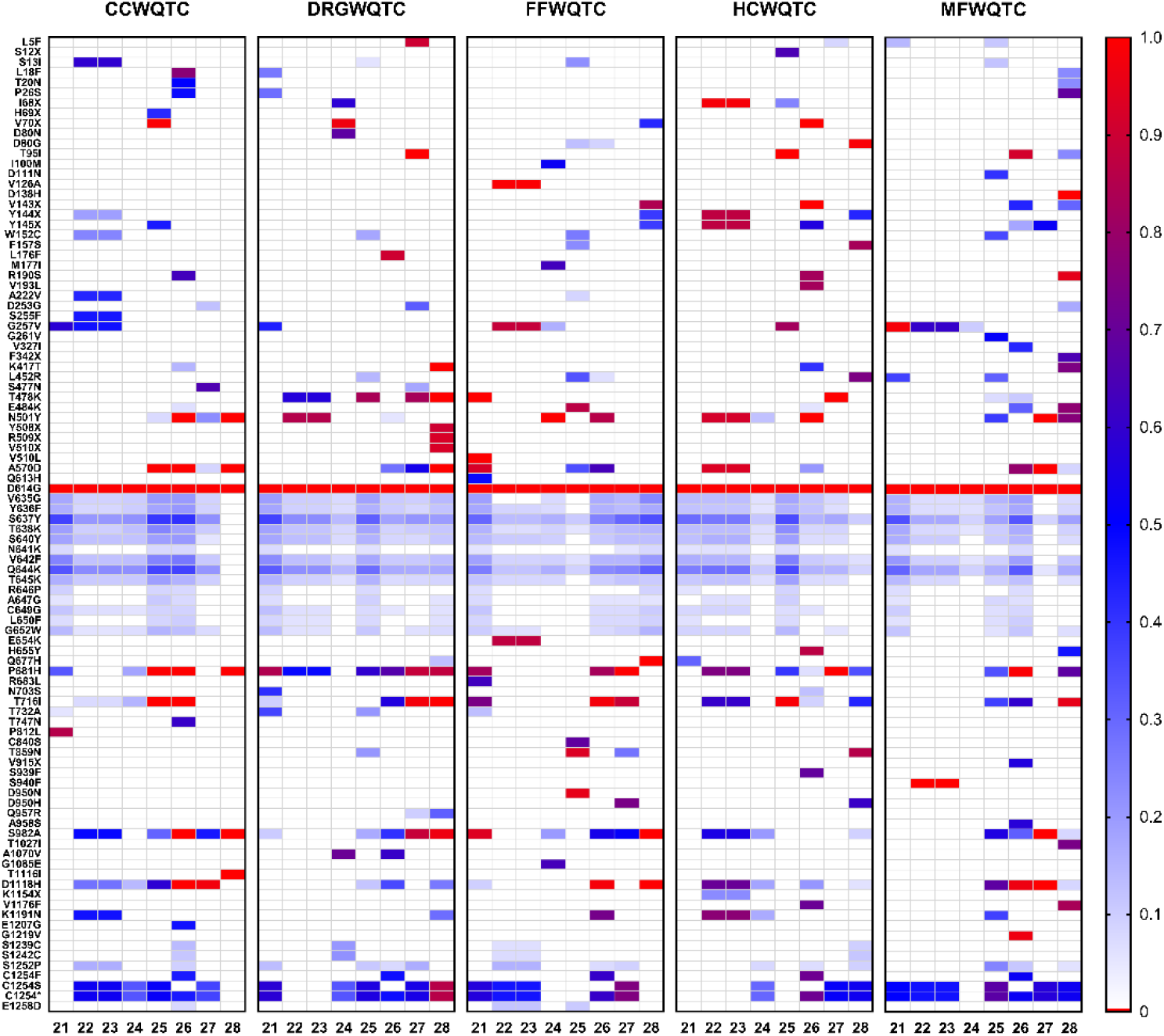
S Protein Polymorphisms Detected in Aggregate Wastewater Over an 8 Week Period. Above are heatmaps for each of the individual wastewater treatment facilities that service the Louisville metropolitan area. Time in weeks is listed on the horizontal axis. Individual polymorphisms that met or exceeded the threshold of 5% prevalence are listed on the left axis. Prevalence, as determined by the number of reads associated with a given polymorphism over total is indicated above in the figure. Silent mutations are not shown.

The most notable dominant polymorphism is the D614G mutation, which was universally detected, as was expected as this mutant lineage is currently dominant globally. Additionally, several other polymorphisms not associated with known VOI/Cs exhibit prolonged detection and significant prevalence-these include a pair of polymorphisms associated with the C-terminal region of the S protein, C1254S and C1254*. Curiously, these mutations are located in the cytoplasmic tail of the S2 fragment of the S protein, and the functions and potential importance of a stop codon in this region are discussed later. In addition to the appearance of polymorphisms associated with the aforementioned VOI/Cs, there is one region of the SARS-CoV-2 S protein that exhibit prolonged detection with moderate levels of sequence prevalence. These include the polymorphisms spanning the SD1 domain of the S1 component of the SARS-CoV-2 S protein. As with the C1254 polymorphisms, the implications of these mutants are discussed below.

## 3. Discussion

### 3.1. Aggregate Wastewater Analyses Are Indicative of Prevalence / Genetic Variation on a Community-Level

In this study we report the quantitative detection of SARS-CoV-2 viral nucleic acid from aggregated wastewater samples in a metropolitan setting of approximately 1 million individuals. As reported in Fig. 1, the detection of SARS-CoV-2 nucleic acid in the aggregated wastewater of Louisville, KY closely mirrored the observed public health data for the corresponding location and time period. Interestingly, a significant lag was observed between the decrease in confirmed cases and the corresponding decrease in SARS-CoV-2 in the aggregate wastewater. Several lines of evidence indicate that this lag is not due to a low rate of exchange in the aggregate setting, as smaller wastewater tributaries exhibited similar phenomena. Moreover, genetic analyses reveal that while the overall levels of SARS-CoV-2 nucleic acid did not appreciably change, the genetic diversity of the SARS-CoV-2 polymorphisms in the wastewater varied to a great extent. Collectively, these observations support the conclusion that the public health data failed to account for all current cases during the period of weeks 14 to 26, and that SARS-CoV-2 prevalence likely remained highly similar to that reported during the peak of the infection. The precise reasons underlying the decrease in the number of confirmed cases detected during this period in time are unknown. One potential reason is a generalized scale back of testing efforts in the Louisville community, which resulted in decreased detection / reporting as asymptomatic individuals were unlikely to seek out testing if not readily available or clinically ill. It should be noted that this period of time corresponds to roughly when large scale vaccination efforts started in Louisville KY, but the speed and magnitude of the decrease is in excess of the vaccination rate during this time. Regardless, altogether, these data reported here confirm the utility and versatility of assessing aggregated wastewater sources for the presence of SARS-CoV-2 nucleic acid.

In addition to readily being able to monitor the disease prevalence within a community, aggregate wastewater can be assessed using next-generation sequencing for the detection of polymorphisms [18,19,21-24]. As shown in Figs. 3-5 the assessment of viral nucleic acid extracted from aggregated wastewater samples is capable of indicating the presence of variants of concern. When comparing aggregate sewersheds to samples of smaller wastewater tributaries there seems to be a lag in the aggregate setting related to resolution likely related to dilution of the virus, however; the virus is generally resolved within a couple weeks as it spreads within the community. Depending on the public health goals and the planned response the total number of samples needed to cover a community can vary, for an observational program the use of aggregated wastewater collected from regional treatment centers should be adequate, but for an early response program to mitigate spread a sewershed level surveillance program may increase resolution and reaction times. Importantly, as shown in Fig. 4, relative changes in strain prevalence can be readily detected, and the emergence of variants within a community can be monitored in real time using aggregated wastewater analyses. Obtaining this information can be informative to the public health response by aiding in general response preparations; however, as the precision with which aggregated wastewater can identify specific individuals infected with particular variants is, obviously, nil, the continued analyses of clinical specimens is warranted.

### 3.2. Sequence Analysis of SARS-CoV-2 Genetic Diversity Reveals Potential Biological Insights

As discussed above, aggregate wastewater can be readily assessed using polymorphism markers associated with VOI/Cs to determine the presence and relative prevalence of specific variants within a community. However, limiting the assessment of sequence diversity to the known VOI/Cs limits the potential of wastewater analyses by seeking to quantify specific markers ignores the primary utility of the next-generation sequencing approach. Wholistic examination of the sequences obtained from the sequencing of aggregate wastewater has the potential to reveal biological insight into the genetic changes driving the continued emergence of SARS-CoV-2. For the purpose of this study, we have limited the region of specific interest to the S protein of SARS-CoV-2. The data in Fig. 6 reveals two interesting phenomena worth further discussion. First, is the apparent significant sequence variation in the SD2 domain of the S1 fragment. This region of S1 is associated with the flexibility with which the Receptor Binding Domain (RBD) can transition between the “up” and “down” conformations, which has been previously associated with receptor binding and entry [27-29]. Curiously, none of the polymorphisms associated with the SD2 domain (which encompasses amino acids 589-677) achieve high prevalence, indicating that perhaps the cost benefit of these variations is not significantly capable of driving selective pressure. It is worth noting that many of these polymorphisms are not currently associated with any VOI/Cs and are relatively uncommon in the SARS-CoV-2 sequence repositories and have not been previously reported in the state of Kentucky. The second is the C1254* mutation which introduces a premature stop codon resulting in the C-terminal truncation of the S protein by 19 amino acids. This particular mutation was amongst the more prevalent and consistently identified polymorphisms in the Louisville metropolitan area, with a prevalence rate of approximately 60% across several weeks. Interestingly, the deletion of the C-terminal 19 amino acids has the potential to interfere with particle assembly, as the C-terminal domain includes a dibasic motif (KxHxx) involved in ER membrane retrieval via an interaction with COPI [30]. This region has been demonstrated to be important to particle assembly in the related betacoronavirus SARS-CoV-1. Thus, the apparent partial selection of a variant with potentially reduced particle production is interesting and may have ramifications on transmission and pathogenesis.

Any discussion of SARS-CoV-2 polymorphism analysis in a complex sample mixture would be remiss if the disclaimer that the sequence diversity observed in wastewater is driven by several factors. One such factor is clearly the number of individuals infected with a particular variant at any given time. Yet another equally important factor is the degree to which particular strains may be shed into wastewater. Thus, the use of wastewater, aggregate or not, to determine the genetic diversity of SARS-CoV-2 in a community is likely not providing a complete understanding of the diversity and selective pressures within a community.

## 4. Conclusions

To conclude, the analysis of aggregated wastewater has proven capable of quantitatively assessing the level of SARS-CoV-2 virus within a community and is often capable of predicting public health trends. In addition to quantitative assessment of SARS-CoV-2 incidence, wastewater surveillance enables the monitoring of genetic diversity at the community-level through the use of polymorphism marker analysis. Furthermore, as evidenced by the data reported here, wastewater surveillance has the potential to identify genetic polymorphisms with potential biological implications without the need for extensive clinical surveillance.

## 5. Materials and Methods

### 5.1. Wastewater Sample Prep

Wastewater samples were collected through a composite sampler with 24-hour composite raw wastewater samples collected into an aseptic 125ml polyethylene terephthalate bottles. Viral particles where precipitated using PEG-8000 (Millipore-Sigma, 89510). Throughout the process, samples were maintained on ice and processed within 12 hrs after collection. For each sample, 40ml of chilled wastewater was passed through a 70 µm cell strainer (VWR, 76327-100) and PEG-8000 and NaCl were added to a final concentration of 12.5 mM and 210 mM, respectively. Samples were refrigerated overnight and then centrifuged at 15,000 x g for 30 mins at 4°C. The pellet was resuspended with 1.1ml TRIzol™ (Thermo Scientific # 15596018) and transferred to a sterile microfuge tube. The TRIzol™ sample was then incubated for 5 mins at room temperature and then centrifuged at 12,000 x g for 5 min at 4°C. The sample was then divided into two 500µl samples, one for isolation and one for archiving at -80°C. The sample for isolation had an additional 500µl of TRIzol added and 900µl of 100% Ethanol. Samples were vortexed and the RNA was isolated using a Direct-zol™ 96 MagBead RNA kit (Zymo Research, R2102) with RNA eluted in 100µl of DNAse/RNAse Free Water. RNA cleanup was done using the Zymo RNA Clean & Concentrator™-5 (Zymo #R1016) according to the manufacturer’s instructions with RNA eluted in 60µl of DNAse/RNAse Free Water. Purified RNA was inspected for yield and quality using a NanoDrop 1000. The number of viral copies in each sample was determined using a probe-based RT-qPCR on a QuantStudio 3 (Applied Biosystems) real-time PCR system using Taq 1-Step Multiplex Master Mix (Thermo Fisher #A28527). The primer and probe sequences are shown in Supplement Table 1 with 5 primer/probe sets used for each sample and all samples ran in triplicate. 4µl of sample was used for each 20µl reaction. PCR cycling conditions were 25°C for 2 min, 50°C for 10 mins, 95°C for 2 min and 45 cycles of 95°C for 2 sec and 60°C for 30 sec. We generated a standard curve for each primer-probe set used and fit the Ct values to extrapolate copies per mL of wastewater. For this publication we are only reporting on the N1 and PMMoV Ct values generated from this methodology. The ΔCt between N1 and PMMoV was used to normalize the N1 concentrations for potential dilution and are graphed over the sampling period.

### 5.2. cDNA Synthesis

The Superscript^®^ IV First-Strand Synthesis System (Thermo Fisher #18091050) was used to generate cDNA with random hexamer primers. The RT reaction was mixed according to manufacturer’s instructions with a final reaction volume of 20 µl and 5 µl of our template RNA added to the mixture. The reverse transcriptase incubation step was performed with sequential incubation at 23°C for 10 min, 50°C for 30 min, and 80°C for 10 min, according to the manufacturer’s protocol with adjustment of the incubation times recommended by Swift Biosciences SNAP low input protocol.

### 5.3. Library Preparation

Libraries were prepared using the Swift Biosciences SNAP low input protocol for SARS-CoV-2 (Swift Bioscience, Ann Arbor, MI, Cat # COSG1V2-96, SN-5X296). 10 µl of cDNA was combined with 20µl of reaction mix and proceeded with multiplex PCR according to protocol. The PCR product was cleaned up using SPRIselect beads (Beckman Coulter, Brea, CA, Cat. No. B23318) at a 1.0X ratio. The purified sample/beads mix was resuspended in 17.4 µl of TE buffer provided in the post-PCR kit. Samples were indexed through PCR with the SNAP Unique Dual Indexing Primers (Swift Bioscience, Ann Arbor, Cat. # SN91096-1-PLATE). The indexing PCR product was further cleaned up and eluted from the beads using a 0.65X PEG NaCl clean-up. The purified libraries were then eluted in 22µL of TE buffer and transferred to fresh tubes and stored at -20°C. 1 additional cycle was added to the multiplex PCR and 2 additional cycles were added to the indexing PCR to obtain higher library yields. Library normalization was performed according to Swift-Bio’s Normalase 2nM final pool protocol. 5 µl of Normalase I Master Mix were added to each 20µl library eluate for a final pool of 2nM and thoroughly mixed. Samples were placed in the thermocycler to incubate at 30°C for 15 min. 5 µl of each library were pooled, and 1µl of Normalase II Master Mix per library was added and thoroughly mixed. The library pool was placed in the thermocycler to incubate at 37°C for 15 min. 0.2 µl of Reagent X1 per library was added to the pool to inactivate Normalase II at 95°C for 2 min and held at 4°C.

### 5.4. Next-Generation Sequencing

Library pool and PhiX were denatured and diluted following Illumina’s directions. Libraries with 1% PhiX spike-in were sequenced at read length 2 × 149 bp using the NextSeq 500/550 Mid Output Kit v2.5 300 Cycles (Illumina, San Diego, CA, Cat# 20024905), targeting 1-5 M reads per library.

### 5.5. Data Analysis

Sequencing reads were analyzed using a custom bioinformatics pipeline. Low quality bases were trimmed using Trimmomatic v0.38 (1), and were then aligned to the NC_045512.2 reference genome using bwa mem v 0.7.17-r1188 (2). Single nucleotide variants (SNVs) relative to the reference were detected using bcftools mpileup (3). SNVs occurring in at least 5% of the reads with at least five separate supporting instances were marked for further interrogation. SNVs occurring at locations of interest as they relate to specific SARS-CoV-2 variants (B.1.1.7, B.1.351, B.1.526, P.1, and B.1.429) were reported for all of the samples (Supplementary Methods Tables 2-5).

## Data Availability

Raw sequence data is available in NCBI Sequence Read Archive under BioProject number PRJNA735936.

## Supplementary Materials

The following are available online at www.mdpi.com/xxx/s1, Table S1: Public Health Information Reported for the City of Louisville, current as of April 2021; and a detailed protocol for the detection of SARS-CoV-2 nucleic acid in wastewater, including all primer sequences.

## Author Contributions

Conceptualization, ECR, SM, KJS, TS, and JF; methodology, ECR, JHC, SW, MZ, DT, KS, IS, MO, and RHH; software, ECR; validation, KS, IS, TS, and JF; formal analysis, ECR, JHC, KJS, and JF; investigation, KS, IS, MP, RHH, KJS, and JF; resources, SW, WZ, DT, SM, KJS, TS, AB, and JF; data curation, ECR, JHC, KS, IS, and RAY; writing—original draft preparation, ECR, JHC, KJS, and JF; writing—review and editing, all authors; visualization, ECR, JHC, KJS, and JF; supervision, ECR, SW, WZ, KJS, and JF; project administration, JF; funding acquisition, TS and AB. All authors have read and agreed to the published version of the manuscript.

## Institutional Review Board Statement

The University of Louisville Institutional Review Board classified this project as Non-Human Subjects Research (NHSR) (reference #: 717950).

## Informed Consent Statement

Not Applicable

## Data Availability Statement

Raw sequence data is available in NCBI’s Sequence Read Archive under BioProject number PRJNA735936.

## Acknowledgments

The following reagent was deposited by the Centers for Disease Control and Prevention and obtained through BEI Resources, NIAID, NIH: Quantitative PCR (qPCR) Control RNA from Heat-Inactivated SARS-Related Coronavirus 2, Isolate USA-WA1/2020, NR 52347.

## References

1. Achak, M.; Alaoui Bakri, S.; Chhiti, Y.; M’Hamdi Alaoui, F.E.; Barka, N.; Boumya, W. SARS-CoV-2 in hospital wastewater during outbreak of COVID-19: A review on detection, survival and disinfection technologies. Sci Total Environ 2021, 761, 143192, doi:10.1016/j.scitotenv.2020.143192.

2. Scott, L.C.; Aubee, A.; Babahaji, L.; Vigil, K.; Tims, S.; Aw, T.G. Targeted wastewater surveillance of SARS-CoV-2 on a university campus for COVID-19 outbreak detection and mitigation. Environ Res 2021, 200, 111374, doi:10.1016/j.envres.2021.111374.

3. Spurbeck, R.R.; Minard-Smith, A.; Catlin, L. Feasibility of neighborhood and building scale wastewater-based genomic epidemiology for pathogen surveillance. Sci Total Environ 2021, 789, 147829, doi:10.1016/j.scitotenv.2021.147829.

4. Fernandez-Cassi, X.; Scheidegger, A.; Banziger, C.; Cariti, F.; Tunas Corzon, A.; Ganesanandamoorthy, P.; Lemaitre, J.C.; Ort, C.; Julian, T.R.; Kohn, T. Wastewater monitoring outperforms case numbers as a tool to track COVID-19 incidence dynamics when test positivity rates are high. Water Res 2021, 200, 117252, doi:10.1016/j.watres.2021.117252.

5. Roka, E.; Khayer, B.; Kis, Z.; Kovacs, L.B.; Schuler, E.; Magyar, N.; Malnasi, T.; Oravecz, O.; Palyi, B.; Pandics, T.; et al. Ahead of the second wave: Early warning for COVID-19 by wastewater surveillance in Hungary. Sci Total Environ 2021, 786, 147398, doi:10.1016/j.scitotenv.2021.147398.

6. Colosi, L.M.; Barry, K.E.; Kotay, S.M.; Porter, M.D.; Poulter, M.D.; Ratliff, C.; Simmons, W.; Steinberg, L.I.; Wilson, D.D.; Morse, R.; et al. Development of Wastewater Pooled Surveillance of Severe Acute Respiratory Syndrome Coronavirus 2 (SARS-CoV-2) from Congregate Living Settings. Appl Environ Microbiol 2021, 87, e0043321, doi:10.1128/AEM.00433-21.

7. Ahmed, W.; Angel, N.; Edson, J.; Bibby, K.; Bivins, A.; O’Brien, J.W.; Choi, P.M.; Kitajima, M.; Simpson, S.L.; Li, J.; et al. First confirmed detection of SARS-CoV-2 in untreated wastewater in Australia: A proof of concept for the wastewater surveillance of COVID-19 in the community. Sci Total Environ 2020, 728, 138764, doi:10.1016/j.scitotenv.2020.138764.

8. Bivins, A.; North, D.; Ahmad, A.; Ahmed, W.; Alm, E.; Been, F.; Bhattacharya, P.; Bijlsma, L.; Boehm, A.B.; Brown, J.; et al. Wastewater-Based Epidemiology: Global Collaborative to Maximize Contributions in the Fight Against COVID-19. Environ Sci Technol 2020, 54, 7754–7757, doi:10.1021/acs.est.0c02388.

9. Li, X.; Kulandaivelu, J.; Zhang, S.; Shi, J.; Sivakumar, M.; Mueller, J.; Luby, S.; Ahmed, W.; Coin, L.; Jiang, G. Data-driven estimation of COVID-19 community prevalence through wastewater-based epidemiology. Sci Total Environ 2021, 789, 147947, doi:10.1016/j.scitotenv.2021.147947.

10. Xu, X.; Zheng, X.; Li, S.; Lam, N.S.; Wang, Y.; Chu, D.K.W.; Poon, L.L.M.; Tun, H.M.; Peiris, M.; Deng, Y.; et al. The first case study of wastewater-based epidemiology of COVID-19 in Hong Kong. Sci Total Environ 2021, 790, 148000, doi:10.1016/j.scitotenv.2021.148000.

11. Hillary, L.S.; Farkas, K.; Maher, K.H.; Lucaci, A.; Thorpe, J.; Distaso, M.A.; Gaze, W.H.; Paterson, S.; Burke, T.; Connor, T.R.; et al. Monitoring SARS-CoV-2 in municipal wastewater to evaluate the success of lockdown measures for controlling COVID-19 in the UK. Water Res 2021, 200, 117214, doi:10.1016/j.watres.2021.117214.

12. Jafferali, M.H.; Khatami, K.; Atasoy, M.; Birgersson, M.; Williams, C.; Cetecioglu, Z. Benchmarking virus concentration methods for quantification of SARS-CoV-2 in raw wastewater. Sci Total Environ 2021, 755, 142939, doi:10.1016/j.scitotenv.2020.142939.

13. Weidhaas, J.; Aanderud, Z.T.; Roper, D.K.; VanDerslice, J.; Gaddis, E.B.; Ostermiller, J.; Hoffman, K.; Jamal, R.; Heck, P.; Zhang, Y.; et al. Correlation of SARS-CoV-2 RNA in wastewater with COVID-19 disease burden in sewersheds. Sci Total Environ 2021, 775, 145790, doi:10.1016/j.scitotenv.2021.145790.

14. Westhaus, S.; Weber, F.A.; Schiwy, S.; Linnemann, V.; Brinkmann, M.; Widera, M.; Greve, C.; Janke, A.; Hollert, H.; Wintgens, T.; et al. Detection of SARS-CoV-2 in raw and treated wastewater in Germany - Suitability for COVID-19 surveillance and potential transmission risks. Sci Total Environ 2021, 751, 141750, doi:10.1016/j.scitotenv.2020.141750.

15. Wu, F.; Zhang, J.; Xiao, A.; Gu, X.; Lee, W.L.; Armas, F.; Kauffman, K.; Hanage, W.; Matus, M.; Ghaeli, N.; et al. SARS-CoV-2 Titers in Wastewater Are Higher than Expected from Clinically Confirmed Cases. mSystems 2020, 5, doi:10.1128/mSystems.00614-20.

16. Gonzalez, R.; Curtis, K.; Bivins, A.; Bibby, K.; Weir, M.H.; Yetka, K.; Thompson, H.; Keeling, D.; Mitchell, J.; Gonzalez, D. COVID-19 surveillance in Southeastern Virginia using wastewater-based epidemiology. Water Res 2020, 186, 116296, doi:10.1016/j.watres.2020.116296.

17. Fuqua, J.L.; Rouchka, E.C.; Waigel, S.; Sokoloski, K.; Chung, D.; Zacharias, W.; Zhang, M.; Chariker, J.; Talley, D.; Santisteban, I.; et al. A rapid assessment of wastewater for genomic surveillance of SARS-CoV-2 variants at sewershed scale in Louisville, KY. medRxiv 2021, 10.1101/2021.03.18.21253604, doi:10.1101/2021.03.18.21253604.

18. Izquierdo-Lara, R.; Elsinga, G.; Heijnen, L.; Munnink, B.B.O.; Schapendonk, C.M.E.; Nieuwenhuijse, D.; Kon, M.; Lu, L.; Aarestrup, F.M.; Lycett, S.; et al. Monitoring SARS-CoV-2 Circulation and Diversity through Community Wastewater Sequencing, the Netherlands and Belgium. Emerg Infect Dis 2021, 27, 1405–1415, doi:10.3201/eid2705.204410.

19. La Rosa, G.; Mancini, P.; Bonanno Ferraro, G.; Veneri, C.; Iaconelli, M.; Lucentini, L.; Bonadonna, L.; Brusaferro, S.; Brandtner, D.; Fasanella, A.; et al. Rapid screening for SARS-CoV-2 variants of concern in clinical and environmental samples using nested RT-PCR assays targeting key mutations of the spike protein. Water Res 2021, 197, 117104, doi:10.1016/j.watres.2021.117104.

20. Bi, C.; Ramos-Mandujano, G.; Tian, Y.; Hala, S.; Xu, J.; Mfarrej, S.; Esteban, C.R.; Delicado, E.N.; Alofi, F.S.; Khogeer, A.; et al. Simultaneous Detection and Mutation Surveillance of SARS-CoV-2 and co-infections of multiple respiratory viruses by Rapid field-deployable sequencing. Med (N Y) 2021, 10.1016/j.medj.2021.03.015, doi:10.1016/j.medj.2021.03.015.

21. Fontenele, R.S.; Kraberger, S.; Hadfield, J.; Driver, E.M.; Bowes, D.; Holland, L.A.; Faleye, T.O.C.; Adhikari, S.; Kumar, R.; Inchausti, R.; et al. High-throughput sequencing of SARS-CoV-2 in wastewater provides insights into circulating variants. medRxiv 2021, 10.1101/2021.01.22.21250320, doi:10.1101/2021.01.22.21250320.

22. Crits-Christoph, A.; Kantor, R.S.; Olm, M.R.; Whitney, O.N.; Al-Shayeb, B.; Lou, Y.C.; Flamholz, A.; Kennedy, L.C.; Greenwald, H.; Hinkle, A.; et al. Genome Sequencing of Sewage Detects Regionally Prevalent SARS-CoV-2 Variants. mBio 2021, 12, doi:10.1128/mBio.02703-20.

23. Martin, J.; Klapsa, D.; Wilton, T.; Zambon, M.; Bentley, E.; Bujaki, E.; Fritzsche, M.; Mate, R.; Majumdar, M. Tracking SARS-CoV-2 in Sewage: Evidence of Changes in Virus Variant Predominance during COVID-19 Pandemic. Viruses 2020, 12, doi:10.3390/v12101144.

24. Nemudryi, A.; Nemudraia, A.; Wiegand, T.; Surya, K.; Buyukyoruk, M.; Cicha, C.; Vanderwood, K.K.; Wilkinson, R.; Wiedenheft, B. Temporal Detection and Phylogenetic Assessment of SARS-CoV-2 in Municipal Wastewater. Cell Rep Med 2020, 1, 100098, doi:10.1016/j.xcrm.2020.100098.

25. Cevik, M.; Tate, M.; Lloyd, O.; Maraolo, A.E.; Schafers, J.; Ho, A. SARS-CoV-2, SARS-CoV, and MERS-CoV viral load dynamics, duration of viral shedding, and infectiousness: a systematic review and meta-analysis. Lancet Microbe 2021, 2, e13–e22, doi:10.1016/S2666-5247(20)30172-5.

26. Yuan, M.; Huang, D.; Lee, C.D.; Wu, N.C.; Jackson, A.M.; Zhu, X.; Liu, H.; Peng, L.; van Gils, M.J.; Sanders, R.W.; et al. Structural and functional ramifications of antigenic drift in recent SARS-CoV-2 variants. Science 2021, 10.1126/science.abh1139, doi:10.1126/science.abh1139.

27. Ou, J.; Zhou, Z.; Dai, R.; Zhang, J.; Zhao, S.; Wu, X.; Lan, W.; Ren, Y.; Cui, L.; Lan, Q.; et al. V367F mutation in SARS-CoV-2 spike RBD emerging during the early transmission phase enhances viral infectivity through increased human ACE2 receptor binding affinity. J Virol 2021, 10.1128/JVI.00617-21, JVI0061721, doi:10.1128/JVI.00617-21.

28. Lan, J.; Ge, J.; Yu, J.; Shan, S.; Zhou, H.; Fan, S.; Zhang, Q.; Shi, X.; Wang, Q.; Zhang, L.; et al. Structure of the SARS-CoV-2 spike receptor-binding domain bound to the ACE2 receptor. Nature 2020, 581, 215–220, doi:10.1038/s41586-020-2180-5.

29. Huang, Y.; Yang, C.; Xu, X.F.; Xu, W.; Liu, S.W. Structural and functional properties of SARS-CoV-2 spike protein: potential antivirus drug development for COVID-19. Acta Pharmacol Sin 2020, 41, 1141–1149, doi:10.1038/s41401-020-0485-4.

30. McBride, C.E.; Li, J.; Machamer, C.E. The cytoplasmic tail of the severe acute respiratory syndrome coronavirus spike protein contains a novel endoplasmic reticulum retrieval signal that binds COPI and promotes interaction with membrane protein. J Virol 2007, 81, 2418–2428, doi:10.1128/JVI.02146-06.

