## Supplementary material for "The Rapid Assessment of Aggregated Wastewater Samples for Genomic Surveillance of SARS-CoV-2 on a City-Wide Scale": Detailed Wastewater Analysis Protocol.docx

**RT-qPCR-based Assay for SARS-COV2 in Kentucky Wastewater**

**Reagents:**

PEG8000 (Sigma Aldrich# 89510)

NaCl (VWR # 0241)

TRIzol™ Reagent (Thermo Scientific # 15596018)

Ethanol (Sigma # E7023)

2-Propanol (Sigma # I9516)

Direct-zol-96 MagBead RNA (Zymo # 2100, VWR# 76211-332)

TaqPath^TM^ 1-Step Multiplex Master Mix 10 mL (Thermo Scientific # A28527)

Micro-Chem Plus (NCL # 0255)

Isopropyl alcohol 70% (VWR # BDH1131)

Zymo RNA Clean & Concentrator (Zymo# R1016)

Primer/probe sets used as shown in Table 2 (Applied Biosystems)

Calf-Guard®, Zoetis, Bovine Rota-Coronavirus Vaccine, Modified Live Virus (Valley Vet #23716)

**Equipment:**

Sterile Cell Strainers 70µm (VWR # 76327-100)

50mL Falcon Tubes (Needs to be rated for 16K x g)

Centrifuge (Thermo Scientific # Sorvall Legend XTR)

Centrifuge (Eppendorf # 5430R)

NanoDrop1000 (Thermo Scientific)

RT-qPCR (ABI #. QuantStudio 3 or 7 System)

Heat-Resistant Films for Real-Time qPCR (VWR #60941-078)

qPCR plates (200 µL VWR # 83007-374 or 100 µL)

**PROTOCOL:**

***Concentration of virus in wastewater samples using PEG8000***

1. Follow BSL Level 2 practices throughout this step (**see safety guidelines below**); Samples should never be opened outside of the Biosafety Cabinet (BSC).
2. Bring the centrifuge rotor, vortex-mixer, buckets of ice, and a beaker containing freshly prepared 10% bleach to the BSC before starting protocol.
3. Spray the exterior of each bottle of sample to be tested (approx. 125 mL) with 2% Micro-Chem, followed by 70% isopropyl alcohol, then place on ice inside the BSC.
4. For each sample to be tested, do the following:

- Label a 50mL high speed centrifuge tube with sample identification information.
- Into one of the labeled tubes, add 4 g PEG and 0.5 g NaCl recap tube.
- Place all prepared tubes in the BSC for filtering.
- Working one sample at a time, place a 70µm cell strainer into each 50mL tube and carefully pipette 40mL of sample into each tube.
- Add 200 µL of BCoV recovery control (See Appendix 1)
- After filtering, securely cap tubes, vortex for 1-2 minutes, or until PEG/NaCl is dissolved, and place them on ice.
- **NOTE:** All used filters and pipettes must be placed in 2% Micro-Chem solution 30 minutes. (samples should always be on ice or at 4^o^C throughout the process).

5. After all samples have been filtered, securely cap the tubes, then place all the tubes into the centrifuge rotor, secure the rotor lid, and incubate the rotor with tubes overnight at 4^o^C.
6. The next morning, turn on the centrifuge to cool it down to 4^o^C and prepare a set of microfuge tubes. You will need one tube per sample, either 1.5 or 2 mL depending on size of pellets.
7. Centrifuge the samples at 15,000 x g for 30 minutes at 4^o^C.

**NOTE:** Do not open the rotor lid unless it is inside the BSC!

8. Working in the BSC, carefully pour the supernatant into a prepared beaker (contains sufficient Micro-Chem to yield a final 5% solution) making sure not to disturb the pellet (gray/black in color).
9. Resuspend the pellet in 1.1 mL TRIzol and transfer this mixture into a sterile microfuge tube.

**NOTE:** Samples can be frozen at this point for later processing if needed.

***RNA extraction using manufacturer’s protocol***

1. Working inside a fume hood, briefly vortex the tubes containing TRIzol™/virus solution, then incubate for 5 mins at RT.
2. Centrifuge the tubes for 5 mins at 12,000 × g at 4°C.
3. Your tubes will now contain three (3) layers: upper layer containing RNA and dirt/debris in the middle/bottom layer.
4. Carefully remove 2x(500µl) the upper layer containing the RNA into two clean, RNAse-free 2ml tubes. **NOTE:** One sample will be used for processing and the other(s) will be used for archival purposes. Store them at -80^o^C.
5. Add an equal volume of TRIzol™ reagent solution (500ul) and add 100% ethanol (900µl) to each processing tube and vortex for 2s.

**NOTE:** This step onwards uses reagents provided in the Zymo RNA isolation kit.

6. Add 15µL MagBinding beads and rock for 10 mins.
7. Transfer the tube to the magnetic stand until beads have pelleted, then aspirate and discard the cleared supernatant.
8. Add 500µL RNA Wash Buffer 1 and mix well. Pellet the beads and discard the supernatant.
9. Add 500µL RNA Wash Buffer 2 and mix well. Pellet the beads and discard the supernatant.
10. Add 500µL 100% ethanol and mix well. Pellet the beads and discard the supernatant.
11. Repeat step 10 (ethanol wash).
12. Dry the beads for 10 minutes in fume hood.
13. To elute RNA from the beads, add 100μL DNase/RNase-Free water and let it stand for 5 minutes.
14. Transfer the tube to the magnetic stand until beads have pelleted, then carefully transfer supernatant to a clean sterile 1.5 ml tube.
15. Add 2 volumes (200 μl) RNA binding buffer and vortex briefly to mix.
16. Add equal volumes (250 μl) of 95 – 100 % ethanol and vortex briefly to mix.
17. Transfer the sample to a Zymo-Spin™ IC Column in a Collection Tube and centrifuge for 30 seconds (12,000 x g). Discard the flow-through.
18. Add 400 μl RNA Prep Buffer to the column and centrifuge for 30 seconds (12,000 x g). Discard

the flow-through.

19. Add 700 μl RNA Wash Buffer to the column and centrifuge for 2 minutes (12,000 x g). Transfer the column carefully into an RNase free tube. Discard the flow-through.
20. To elute the RNA, add 62 μl of RNase-Free Water (provided) to the center of the

white filter membrane. Incubate for 1 min at room temperature and centrifuge at 10,000 x g for 1 min at room temperature.

21. Add eluate to the filter membrane back again and centrifuge at 10, 000 x g for 2 min at room temperature.
22. Discard the Zymo-Spin™ IC Column. The RNA is now ready for downstream applications.

**NOTE:** Samples can be stored at -80°C at this point.

***1-Step RT-qPCR***

1. Use a NanoDrop1000 (Thermo Scientific), or similar spectrophotometer, to measure the yield and quality of the eluted RNA samples.
2. Pipet 4µL of each RNA sample into triplicate wells, being sure to use a RT-qPCR machine compatible 96-well plate.
3. Prepare your PCR Reaction Master Mix as shown in Table 1.
4. The multiplex PCR is designed using three (3) primer/probe sets. Set 1 includes primers/probes for N1. Set 2 includes primer/probes for BCoV. Set 3 includes primers/probes PMMoV and CrAssphage (See Table 2).

**NOTE:** Each plate should be set up to include corresponding positive and negative controls.

5. Centrifuge the plate for 2 min at 1200 × g.
6. RT-qPCR conditions are shown in Table 3.

***Analysis***

- Analyze samples using C_q_ value of standards as shown in table 5.
- The no template control sample should have no C_q_ value or a value above 36.
- Positive controls should show less than 20% variance from the mean.
- A rolling average of PMMoV will be calculated for each site and the weekly difference will be used to normalize the data for fecal input.
- BCoV is spiked into the raw sample and its recovery calculated based upon the C_q_ value. The recovery is used to normalize the data set for viral recovery.

***Safety Guidelines***

Procedures that concentrate viruses, such as precipitation or membrane filtration, should be performed in a BSL-2 laboratory with unidirectional airflow and BSL-3 precautions, including respiratory protection and a designated area for donning and doffing PPE. The donning and doffing space should not be in the workspace. Work should be performed in a certified Class II BSC.

Reference: <https://www.cdc.gov/coronavirus/2019-nCoV/lab/lab-biosafety-guidelines.html>

**Table 1**

| **Component** | **Volume for 20μL reaction** | **Final Concentration** |
| --- | --- | --- |
| TaqPath^TM^ 1-Step Multiplex Master Mix (4X) | 5µL | 1 X |
| Forward primer(s) | 0.1µL | 500nM |
| Revesre primer(s) | 0.1µL | 500nM |
| Probe(s) | 0.04µL | 125nM |
| RNA Template | 4µL | Variable |
| Nuclease-free water | 10.76µL | Variable |

**Table 2**

| **Primer Name** | **Sequence** | **Probes** | **Final Conc.** |
| --- | --- | --- | --- |
| 2019-nCoV_N1-F | 5’-GACCCCAAAATCAGCGAAAT-3’ | None | 500nM |
| 2019-nCoV_N1-R | 5’-TCTGGTTACTGCCAGTTGAATCTG-3’ | None | 500nM |
| 2019-nCoV_N1-P | 5’-**FAM**-ACCCCGCATTACGTTTGGTGGACC-**QSY**-3’ | FAM, QSY | 125nM |
| BCoV-F | CTGGAAGTTGGTGGAGTT | None | 500nM |
| BCoV-R | ATTATCGGCCTAACATACATC | None | 500nM |
| BCoV-P | 5’-**VIC**-CCTTCATATCTATACACATCAAGTTGTT -**QSY**-3’ | JUN, QSY | 125nM |
| PMMoV-F | 5’GAGTGGTTTGACCTTAACGTTTGA-3’ | None | 500nM |
| PMMoV-R | 5’-TTGTCGGTTGCAATGCAAGT-3’ | None | 500nM |
| PMMoV-P | 5’-**VIC**-CCTACCGAAGCAAATG-**QSY**-3’ | VIC, QSY | 125nM |
| CrAssphage-F | 5’-CAGAAGTACAAACTCCTAAAAAACGTAGAG-3’ | None | 500nM |
| CrAssphage-R | 5’-GATGACCAATAAACAAGCCATTAGC-3’ | None | 500nM |
| CrAssphage-P | 5’-**JUN**-AATAACGATTTACGTGATGTAAC-**QSY**-3’ | JUN, QSY | 125nM |

**Probe listing for Ordering**

| 2019-nCoV_N1-P | ACCCCGCATTACGTTTGGTGGACC | 20k pmol | HPLC | liquid | FAM | QSY |
| --- | --- | --- | --- | --- | --- | --- |
| BCoV-P | CCTTCATATCTATACACATCAAGTTGTT | 20k pmol | HPLC | liquid | JUN | QSY |
| PMMoV-P | CCTACCGAAGCAAATG | 20k pmol | HPLC | liquid | VIC | QSY |
| CrAssphage-P | AATAACGATTTACGTGATGTAAC | 20k pmol | HPLC | liquid | JUN | QSY |

**Table 3**

| **Step** | **Stage** | **Cycles** | **Temperature** | **Time (Fast)** |
| --- | --- | --- | --- | --- |
| UNG incubation | 1 | 1 | 25°C | 2 mins |
| Reverse Transcription | 2 | 1 | 53°C | 10 mins |
| Polymerase Activation | 3 | 1 | 95°C | 2 mins |
| Amplification | 4 | 45 | 95°C | 5 s |
|  |  |  | 60°C | 30 s |

**Table 4 Positive Controls**

| **Control Type** | **Against** | **Source** |
| --- | --- | --- |
| Negative | Background | Tap water |
| Positive | CoV Nucleocapsid (N1) | Heat-Inactivated SARS-CoV-2, Isolate USA-WA1/2020 |
| Positive | Bovine CoV | Calf-Guard®, Zoetis  Bovine Rota-Coronavirus Vaccine  Modified Live Virus |
| Positive | Pepper Mild Mottle Virus | Plasmid |
| Positive | Cross-assembly phage | Plasmid |

**Table 5 Linear Epitope Standards Primer F, Primer R, Probe**

| **Target** | **Linear Epitope** |
| --- | --- |
| CoV Nucleocapsid (N1) | TTGCCAGGAACCTAAATTGGGTAGTCTTGTAGTGCGTTGTTCGTTCTATGAAGACTTTTTAGAGTATCATGACGTTCGTGTTGTTTTAGATTTCATCTAAACGAACAAACTAAAATGTCTGATAATGGACCCCAAAATCAGCGAAATGCACCCCGCATTACGTTTGGTGGACCCTCAGATTCAACTGGCAGTAACCAGAATGGAGAACGCAGTGGGGCGCGATCAAAACAACGTCGGCCCCAAGGTTTACCCAATAATACTGCGTCTTGGTTCACCGCTCTCACTCAACATGGCAAGGAAGAC |
| Bovine CoV | TATCATCTTAACTATTTTCAATTGCGTGTATGCGTTGAATAATGTGTATCTTGGCTTCTCTATAGTTTTTACTATAGTGGCCATTATCTTGTGGATTGTGTATTTTGTGAATAGTATCAGGTTGTTTATTAGAACTGGAAGTTGGTGGAGTTTCAACCCAGAAACAAACAACTTGATGTGTATAGATATGAAGGGAAGGATGTATGTTAGGCCGATAATTGAGGACTACCATACCCTTACGGTCACAATAATACGTGGTCATCTTTACATGCAAGGTATAAAACTAGGTACTGGCTATTCTTTGTCAGATTTGCCAGCTTATGTGACTGTTGCTAAGGTCTCGCACCTGCTCACGTATAA |
| Pepper Mild Mottle Virus | TACAATGCTTTGTCAGAAATCTCAATTCTTAAAGACAGTGACAAGTTTGATGTTGATGTTTTTTCCCGGATGTGTAATACATTAGGCGTAGATCCATTGGTGGCAGCAAAGGTAATGGTAGCTGTGGTTTCAAATGAGAGTGGTTTGACCTTAACGTTTGAGAGGCCTACCGAAGCAAATGTCGCACTTGCATTGCAACCGACAATTACATCAAAGGAGGAAGGTTCGTTGAAGATTGTGTCGTCAGACGTAGGTGAGTCCTCAATCAAGGAAGTGGTTCGAAAATCAGAGATTTCTATGCTTGGTCTAACAGGCAACACAGTGTCCGATGAGTTCCAAAGAAGTACAGAAATCGAGTCG |
| Cross-assembly phage | CATAGAGCTGCATTGTTAGTATTGCTAATGGTGCAGCTCTTATTTTATCTAATAATTCTTTTAATTACTTTAATTATGTCGACAGAAAAAGAAATTAAGAATGAAGCTACTGTTGTAGCAAGTGCTGAACAAACTGCTAATGCAGAAGTACAAACTCCTAAAAAACGTAGAGGTAGAGGTATTAATAACGATTTACGTGATGTAACTCGTAAAAAGTTTGATGAACATACTGATTGTAATAAAGCTAATGGCTTGTTTATTGGTCATCTTGAAGATGTTAAAGTTGATTGGGCTACATTGAAAGATGATGTTCAAGGTATGCCTTCATTTGCTGGTATGAGTATTCCTTATCTTACATTTACTTTTGCTAGTAATCATGAAAATATCAATGAACGTCGTTATGTAACTCAACGTCTTCTT |

**Appendix 1 – Making BCoV Recovery Control**

1. Calf-Guard®, Zoetis, Bovine Rota-Coronavirus Vaccine, Modified Live Virus (Valley Vet #23716). Pull one vial of freeze-dried vaccine and one vial of sterile diluent.
2. Aseptically rehydrate the freeze-dried vaccine with 3 mLs of the provided sterile diluent. Vortex briefly to resuspend.
3. In a sterile RNase DNase free bottle add 247.5 mL of RNase/DNase free water. To the same bottle add 2.5 mL of the rehydrated vaccine and mix well.
4. Aliquot in 1 mL volumes in 1.5 mL RNase/DNase free tubes. Freeze at -20°C. Do not freeze-thaw.
